# Pathogen Control System for Buildings

**DOI:** 10.64898/2025.12.02.25341488

**Authors:** Samuel Fernandes, Raja Sengupta

## Abstract

**Background:** Environmental control systems in buildings are typically designed to maintain occupant comfort while minimizing energy use. However, the significant role of airborne pathogens in respiratory illness transmission has highlighted the imperative to address how these control systems can mitigate infection risk. Traditional CO_2_-based ventilation control does not necessarily correlate with infectious aerosol presence, limiting its effectiveness for pathogen mitigation.

**Objective:** To develop and evaluate a pathogen control system (PCS) that combines real-time pathogen sensing with in-duct germicidal ultraviolet (GUV) irradiation to reduce infection risk while maintaining energy efficiency and occupant comfort.

**Methods:** We developed a closed-loop control system using pathogen air quality (PAQ) sensors with hysteretic threshold control (7-20 copies/m^3^) to dynamically activate GUV systems achieving 99% single-pass inactivation efficiency. System performance was evaluated across four activity scenarios (1.33-750 copies/s generation rates) in a simulated 70 m^3^ office environment using eight complementary metrics: peak concentration (*C*_peak_), steady-state concentration (*C*_ss_), clearance time improvement (Δ*t*_clear_), time to safety (*t*_safe_), cumulative inhaled dose (*D*_*inh*_), infection risk probability (*P*_risk_), equivalent clean air rate (ECAi), and energy consumption.

**Results:** In talking scenarios, the PCS reduced peak concentration from 40 to 22 copies/m^3^ (45% reduction), time to safety from 75 to 25 min (67% improvement), cumulative inhaled dose from 1.1×10^−2^ to 4.4×10^−3^ copies (60% reduction), and infection risk from 56.07% to 28.88%. In high activity scenarios, peaks decreased from 90 to 45 copies/m^3^, time to safety from 90 to 30 min, dose from 2.4×10^−2^ to 8.4×10^−3^ copies (65% reduction), and risk from 83.93% to 47.83% (43% relative reduction). Baseline active control increased ECAi from 108 to 191 m^3^/h, with geometric scaling enabling pathways to full ASHRAE 241 compliance (920-3,680 m^3^/h). System performance was robust across sampling intervals (30-300 s) while achieving 37-52% energy savings through duty-cycle operation.

**Significance:** This study provides the first comprehensive quantitative framework for sensor-based pathogen control in building environments. The demonstrated ability to achieve substantial infection risk reduction while maintaining energy efficiency supports the viability of pathogen-responsive building control as an effective intervention for indoor air quality management. Results establish fundamental design principles and performance benchmarks that can inform regulatory guidelines, building codes, and public health recommendations for pathogen control system deployment in the era of healthy buildings.

**Impact Statement:** This research addresses a critical gap in building environmental control by demonstrating how real-time pathogen sensing can enable targeted, energy-efficient disinfection strategies that traditional CO_2_-based systems cannot achieve. By providing quantitative evidence that sensor-based pathogen control systems can reduce infection risk by 43-67% across realistic occupancy scenarios while maintaining operational efficiency, this work establishes a scientific foundation for next-generation healthy building technologies. The systematic evaluation framework and performance benchmarks developed herein directly support evidence-ng based implementation of pathogen-responsive building control systems, contributing to improved occupant health outcomes and enhanced pandemic preparedness in built environments. These findings are particularly relevant for the Journal of Exposure Science and Environmental Epidemiology’s focus on environmental health and exposure assessment, as they provide quantitative tools for evaluating and optimizing indoor air quality interventions that reduce infectious disease transmission risk.

## 1 Introduction

In this paper we present a pathogen control system (PCS) that combines a real-time pathogen sensor[1], with in-duct GUV irradiation and ventilation to reduce the risk of infection. Our control system seeks to minimize infection risk quickly, while balancing energy costs and maintaining comfort. Feedback control is well established in buildings, however, these closed loop controls mainly focus on optimizing comfort and energy[2]–[8]. Recent efforts to control disease transmission through ventilation have utilized CO_2_ sensors [9]. However, the development of real-time pathogen sensing has opened the possibility of developing feedback control for infection risk, which is what we explore in this paper. As a practical example, we examine a single-zone system and demonstrate through simulation that the controller effectively controls pathogen levels while maintaining energy efficiency and comfort.

### 1.1 Background and Motivation

Environmental control systems in buildings are essential for maintaining a comfortable and safe indoor environment while optimizing energy efficiency [10], [11]. However, these systems have become increasingly important given the critical role that airborne pathogen transmissions have played in various respiratory epidemics, such as influenza, severe acute respiratory syndrome, and Middle East respiratory syndrome [12]–[15]. Factors like environmental conditions, pathogen characteristics, and human activities can impact the airborne survival and transmission of these infectious pathogens [13], [16], [17]. Typically, HVAC control strategies, such as occupancy-based ventilation, aren’t designed to adapt to the dynamics of airborne infection transmission, except in specific environments like hospital isolation rooms [18], [19]. The recent ASHRAE Standard 241 aims to broaden infection control to encompass a wider range of buildings, offering guidance for enhancing indoor air quality and decreasing the propagation of airborne pathogens[20].

Environmental control strategies to mitigate airborne infection risks in buildings can broadly be categorized into three approaches: ventilation, filtration, and disinfection. This categorization is primarily based on physical mechanisms for removing infectious agents from the air [21]–[31]. Natural or mechanical ventilation can effectively dilute the concentration of infectious airborne contaminants by introducing fresh outdoor air, but increased ventilation is often impractical due to HVAC system constraints and energy costs [32], [33]. Additionally, improper HVAC system implementation of ventilation can potentially exacerbate the transmission of airborne illnesses [5]–[8], [34]. Filtration, using high-efficiency particulate air filters, can remove a significant portion of airborne contaminants, but this approach is limited by filter effectiveness against smaller viral particles and the need for regular maintenance [35]–[39]. Disinfection, primarily achieved through germicidal ultraviolet (GUV) irradiation, uses short-wave UV energy to inactivate viruses, bacteria, and fungi by forming photodimers in nucleic acids, preventing transcription and replication [24], [40]–[44]. GUV irradiation has been found to be a highly effective method for disinfecting large volumes of indoor air and for inactivating a wide variety of microbial pathogens, including protozoa, fungi, bacteria, and viruses[45]–[50]. The required UV dose may vary depending on the specific pathogen, for instance fungal spores are among the most resistant microorganisms, but GUV has been shown to be effective in reducing mold growth on air conditioning coils and drip pans [51]–[53]. Compared to mechanical ventilation and portable air cleaners, GUV with well-mixed room air has been found to provide the equivalent of up to 24 air changes per hour under real-world conditions [53]. The mechanism is facilitated by the well-mixed nature of the room air, which is achieved through several factors [54], [55]. Firstly, convective air currents generated by occupant body heat effectively mix the air between the upper and lower zones of the room. Occupant movement, ventilation diffusers, and supply air temperature gradients also contribute to the thorough mixing of the room air [56]. Additionally, low-velocity ceiling fans can further enhance this mixing process in a quiet and cost-effective manner. In buildings with inefficient mechanical ventilation systems, especially under high-risk conditions, GUV may be the only practical method for controlling airborne infectious agents [57], [58].

Recent research has highlighted the use of CO_2_ to estimate infection risk [59]–[62]. However, CO_2_ levels do not necessarily correlate with the presence of infectious aerosols. Therefore relying solely on CO_2_-based ventilation control to minimize infection risk can lead to unnecessary energy costs. The recent development of pathogen sensors, such as the pathogen air quality (PAQ) monitor and others [1], [63]–[65], shows considerable promise for real-time monitoring of airborne pathogens. Laboratory experiments demonstrate that the PAQ monitor for real-time direct detection of SARS-CoV-2 aerosols has a device sensitivity of 77–83 and a limit of detection of 7-35 viral RNA copies/m^3^ of air. The PAQ monitor is suitable for point-of-need surveillance of SARS-CoV-2 variants in indoor environments and can be adapted for multiplexed detection of other respiratory pathogens of interest [66]–[74].

### 1.2 Research Questions and Paper Organization

Despite progress in pathogen detection and disinfection, there is a lack of research on integrating them into closed-loop building control systems, which is what we explore in this paper. We address questions regarding the control efficacy, activity-dependent performance, sensor sampling frequency, GUV system design, actuator efficiency and regulatory compliance of the PCS for infection risk mitigation. Specifically we address the following questions:

- What is the quantitative reduction in infection risk (%) achieved by the sensor-based control system compared to uncontrolled conditions, and how do peak concentration (*C*_peak_), steady-state concentration (*C*_ss_), and clearance time improvement (Δ*t*_clear_) change under controlled vs. uncontrolled scenarios?
- How does controller performance vary across different human activities with varying pathogen generation rates (*G*_*i*_, where *i* ∈ {sitting, talking, exercising, coughing})?
- How does sensor sampling interval (Δ*t*_sample_) affect actuator response time and overall system performance?
- How do geometric modifications to UVGI exposure chambers (duct length and cross sectional area) and multi-unit installations quantitatively affect pathogen inactivation efficiency, and what is the functional relationship between UVGI efficiency (*η*) and infection risk reduction?
- How does the system performance compare against ASHRAE 241 Equivalent Clean Air rate (ECAi) requirements?

This paper is organized as follows: Section 2 establishes the mathematical and control system framework needed to evaluate pathogen control efficacy and system performance; Section 3 describes the experimental setup to test the performance of the PCS in a simulated environment ; Section 4 presents results addressing control effectiveness, activity-dependent performance, and ASHRAE 241 compliance; Section 5 discusses optimization strategies and implementation implications; and Section 6 concludes with synthesis of findings and future research directions.

## 2 Methods

### 2.1 Mathematical Framework

The PCS is governed by fundamental mass balance equations that describe the concentration dynamics of airborne pathogens in indoor spaces. The system behavior is modeled using:

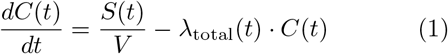

where *C*(*t*) is the pathogen concentration (copies/m^3^), *S*(*t*) is the generation rate (copies/s), *V* is the room volume (m^3^), and *λ*_total_(*t*) is the total removal rate (1/s). The total removal rate combines ventilation and UVGI effects:

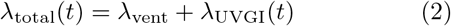

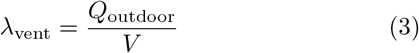

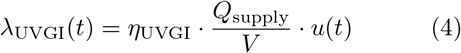

where *Q*_outdoor_ is the outdoor air volumetric flow rate (m^3^/s), *η*_UVGI_ = 0.99 is the UV inactivation efficiency, *Q*_supply_ is the supply airflow rate, and *u*(*t*) is the controller state (0 or 1).

### 2.2 System Architecture

The PCS employs a closed-loop architecture that continuously monitors indoor pathogen concentrations and dynamically adjusts disinfection strategies (Figure 1). The control loop receives real-time concentration data from a PAQ sensor as its primary input, processing this information through a threshold based controller that determines UVGI activation states. The system inputs include: (1) continuous pathogen concentration measurements from the PAQ sensor, (2) configurable control thresholds for UVGI activation/deactivation, and (3) ventilation parameters including outdoor air fraction and total supply flow rate. The system outputs consist of binary UVGI on/off commands and maintains constant ventilation parameters throughout operation, thereby not impacting occupant comfort.

**Figure 1.**
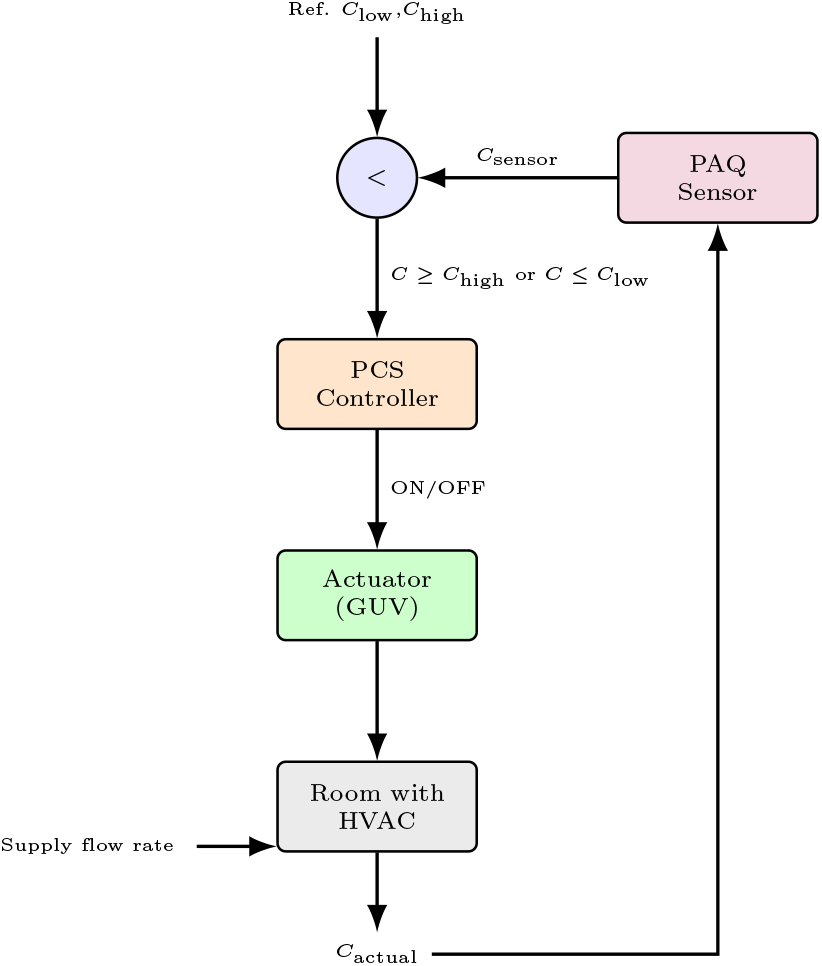
Pathogen control system architecture showing the PCS controllers with concentration sensor feedback, based on the control system design schematic.

### 2.3 Control Strategy

The system implements a hysteretic threshold based control algorithm with dual concentration thresholds to prevent excessive cycling while maintaining acceptable risk levels. The threshold values were selected based on quanta emission rate data and dose-response relationships established in recent literature [75], [76], but can be adapted. The control law is mathematically expressed as:

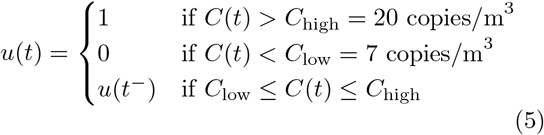

where *u*(*t*) is the UVGI control signal (1 = ON, 0 = OFF), *C*(*t*) is the measured pathogen concentration, and *u*(*t*^−^) represents the previous control state. The controller activates UVGI when pathogen concentration exceeds 20 copies/m^3^ and deactivates when concentration falls below 7 copies/m^3^, creating a 13 copies/m^3^ deadband that reduces switching frequency and extends equipment life. The control hierarchy prioritizes UVGI activation over ventilation adjustments due to superior single-pass efficiency (90% vs. 30% dilution from outdoor air) and faster response dynamics. Ventilation parameters remain constant to maintain thermal comfort and ASHRAE compliance, while UVGI provides rapid, targeted pathogen inactivation without affecting temperature or humidity setpoints. The system does not explicitly model energy-comfort-risk trade-offs, focusing primarily on pathogen mitigation effectiveness.

### 2.4 Sensor model

The PAQ sensor simulation, models a continuous monitoring sensor with configurable sampling intervals ranging from 1 to 300 seconds, with a default of 120 seconds for optimal balance between response time and measurement stability. Real sensors exhibit non-ideal characteristics including finite sensitivity and sampling delays. The model incorporates a sensitivity threshold of 85% detection probability, sampling rates of 30 seconds, 2 minutes, and 5 minutes, as well as measurement noise and detection delays. While the specific detection mechanism is abstracted in the simulation, the sensor is assumed to quantify airborne pathogen concentrations in real-time. The sensor response is modeled as an exponential rise function:

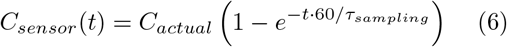

where *τ*_*sampling*_ represents the sampling interval. This model captures the inherent lag between actual pathogen release and sensor detection, critical for understanding control system performance during transient events.

### 2.5 UVGI System

The UVGI system operates at a fixed power level achieving 99% single-pass inactivation efficiency for recirculated air. The control system modulates UVGI exposure through binary on/off switching rather than variable power control, optimizing for simplicity and f reliability over continuous dose adjustment. The system’s effectiveness is modeled through a survival fraction parameter:

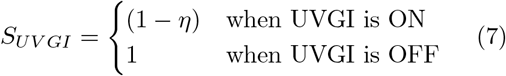

where *η* = 0.99 represents the single-pass efficiency. The UVGI effectively reduces pathogen concentration in recirculated air. The UVGI dose delivery mechanism assumes instantaneous inactivation upon air passage through the UV field, neglecting detailed UVC dose calculations (J/m^2^) in favor of an empirical single-pass efficiency metric. This simplification is justified for in-duct systems where residence time and irradiance levels are well-characterized through prior experimental validation.The system maintains compliance with ventilation standards through fixed minimum OA rates rather than demand-controlled ventilation, ensuring consistent indoor air quality regardless of pathogen detection status. While economizer logic could be integrated, the current implementation prioritizes infection control over energy optimization. Energy consumption of the PCS is calculated as the product of UVGI activation time and power consumption:

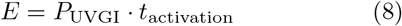

where *P*_UVGI_ is the UVGI system power (kW) and *t*_activation_ is the total time the UVGI system operates during the simulation period (hours).

### 2.6 Infection risk

Real-time infection risk estimation employs a dose-response risk model using an alternative quanta conversion approach. The cumulative inhaled dose is calculated as:

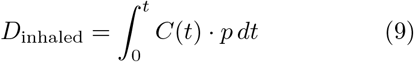

where *D*_inhaled_ is total inhaled dose (copies), *C*(*t*) is time-varying concentration (copies/m^3^), *p* is pulmonary ventilation rate (m^3^/h), and *t* is exposure time (h). This dose is converted to equivalent quanta:

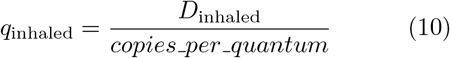

In this context, a “quantum” is defined as the number of inhaled infectious virus-laden aerosols required to infect 63.21% of susceptible persons, based on the Wells-Riley exponential dose-response model [75]. The value *copies per quantum* = 0.0129 in our model represents the conversion factor from pathogen copies to quanta. This value corresponds to approximately ≈ 77.5 RNA copies per quantum (1*/*0.0129 ≈ 77.5), which is calibrated for our specific pathogen based on empirical data and is more conservative than values reported by Aganovic et. al [75] for various SARS-CoV-2 variants.

The infection risk probability then follows:

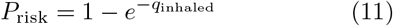

The infection risk model integrates real-time concentration measurements with activity-dependent breathing rates to calculate cumulative pathogen exposure. System performance is evaluated against ASHRAE Standard 241 [20] using the Equivalent Clean Airflow for infection control (ECAi):

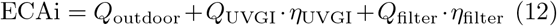

We assume uniform mixing within the zone and do not account for spatial concentration gradients or proximity effects between infectious sources and susceptible occupants. The real-time risk calculation enables predictive control strategies, allowing the system to activate mitigation measures before reaching unacceptable risk thresholds.

## 3 Experiments

We motivate the operation of the PCS with a simulation model of a single-zone space with volume *V* = 70 m^3^, representing a typical office or classroom environment (Figure 2). The simulation includes an infectious occupant (infector) present for the first 15 minutes, generating pathogenic aerosols at rates determined by their activity level (ranging from 1.33 to 750 copies/s). The PCS is integrated with a constant-volume ventilation system operating at *Q*_supply_ = 0.1 m^3^/s (212 CFM) total supply airflow with a default outdoor air fraction of 0.3 (*Q*_outdoor_ = 0.03 m^3^/s or 63.6 CFM, *Q*_recirc_ = 0.07 m^3^/s or 148.3 CFM). The baseline UVGI system in the duct has dimensions of 0.30 m × 0.20 m × 0.10 m, with irradiance of 20 W/m^2^. These components interact through mass balance equations that govern pathogen transport, dilution, and inactivation within the conditioned space, as described in Equations (1-4).

**Figure 2.**
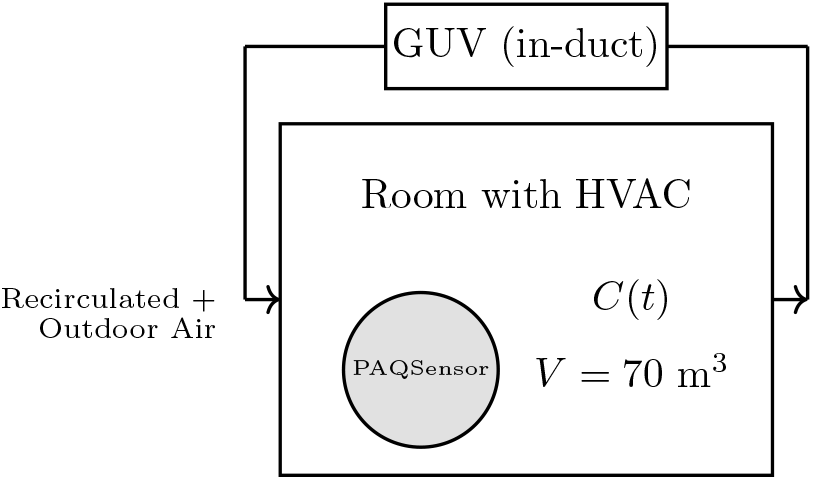
Simplified single-zone experimental setup showing room with HVAC system, in-duct GUV, and PAQ sensor for pathogen concentration monitoring.

### 3.1 Activity Scenarios

Four distinct generation activity scenarios were simulated to represent different pathogen generation rates, ranging from low-level background emissions to acute respiratory events (Table 2).

**Table 1.** Single-Zone Office Space Parameters.

| Parameter | Value |
| --- | --- |
| Room volume | $70 \text{ m}^3$ |
| Supply airflow rate | $0.1 \text{ m}^3/\text{s}$ (212 CFM) |
| Outdoor air fraction | 0.3 (30% outdoor air) |
| Outdoor airflow rate | $0.03 \text{ m}^3/\text{s}$ (63.6 CFM) |
| Duct dimensions | $0.30 \text{ m} \times 0.20 \text{ m} \times 0.10 \text{ m}$ |
| UVGI irradiance | $20 \text{ W}/\text{m}^2$ |

**Table 2.**
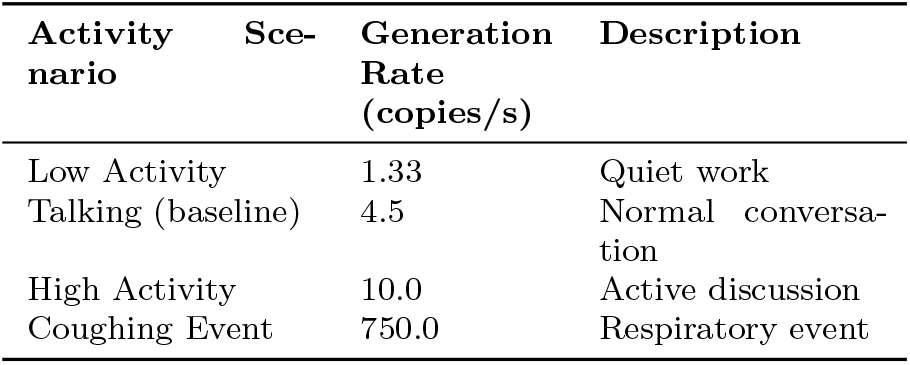
Activity Scenarios and Pathogen Generation Rates.

| Activity Scenario | Generation Rate (copies/s) | Description |
| --- | --- | --- |
| Low Activity | 1.33 | Quiet work |
| Talking (baseline) | 4.5 | Normal conversation |
| High Activity | 10.0 | Active discussion |
| Coughing Event | 750.0 | Respiratory event |

The low activity scenario (1.33 copies/s) represents quiet work environments such as reading, writing, or computer-based tasks with minimal vocalization and reduced respiratory emissions [77], [78]. The talking scenario (4.5 copies/s) models normal conversation levels typical in office meetings, classroom discussions, or collaborative work environments where occupants engage in regular speech patterns [60], [62]. High activity scenarios (10.0 copies/s) encompass active discussions, presentations, or physical activities that increase respiratory rate and pathogen generation through elevated breathing and vocalization [22], [77]. The coughing event scenario (750.0 copies/s) represents acute respiratory events that can dramatically increase airborne pathogen concentrations over short durations, simulating conditions during illness or respiratory distress [12], [13]. These generation rates span nearly three orders of magnitude, enabling comprehensive evaluation of system performance across the full spectrum of realistic indoor emission scenarios [21], [23].

### 3.2 Performance Metrics

System effectiveness is quantified using eight complementary performance metrics—peak pathogen concentration (*C*_peak_), steady-state concentration (*C*_ss_), clearance time improvement (Δ*t*_clear_), time to reach safe levels (*t*_safe_), cumulative inhaled dose (*D*_*inh*_), infection risk probability (*P*_risk_), equivalent clean air rate (ECAi), and energy consumption (*E*)—which collectively characterize transient suppression capacity, sustained removal performance, integrated exposure, health outcome translation, and operational efficiency (Table 3).

**Table 3.** System Performance Metrics (with concise definitions)

| Parameter | Symbol | Concise Definition |
| --- | --- | --- |
| Peak pathogen concentration | $C_{\text{peak}}$ | Maximum concentration during emission |
| Steady-state concentration | $C_{\text{ss}}$ | Average controlled concentration after transients |
| Clearance time improvement | $\Delta t_{\text{clear}}$ | Reduction in time-to-safe compared to no control |
| Time to reach safe levels | $t_{\text{safe}}$ | Time when concentration first reaches the safe threshold after source stops |
| Cumulative inhaled dose | $D_{\text{inh}}$ | Total inhaled pathogen copies over the exposure period |
| Infection risk probability | $P_{\text{risk}}$ | Probability of infection based on the inhaled dose (Wells–Riley) |
| Equivalent clean air rate | ECAi | Total effective clean airflow for infection control |
| Energy consumption | $E$ | UVGI electrical energy use |

**Table 4.** System Performance Metrics for Different Activities.

| Activity | Control | $C_{\text{peak}}$<br>(copies/m<sup>3</sup>) | $C_{\text{ss}}$<br>(copies/m<sup>3</sup>) | $t_{\text{clear}}$<br>(min) | $t_{\text{safe}}$<br>(min) | $D_{\text{inh}}$<br>(copies) | $P_{\text{risk}}$<br>(%) |
| --- | --- | --- | --- | --- | --- | --- | --- |
| Low Activity | No Control | 14.2 | 13.3 | 120.0 | 42.5 | 3.1e-03 | 21.63 |
| Low Activity | PCS Controller not activated | 14.2 | 13.3 | 120.0 | 42.5 | 3.1e-03 | 21.63 |
| Talking | No Control | 40.0 | 38.0 | 120.0 | 75.0 | 1.1e-02 | 56.07 |
| Talking | PCS Control | 22.0 | 20.0 | 60.0 | 25.0 | 4.4e-03 | 28.88 |
| High Activity | No Control | 90.0 | 85.0 | 120.0 | 90.0 | 2.4e-02 | 83.93 |
| High Activity | PCS Control | 45.0 | 40.0 | 45.0 | 30.0 | 8.4e-03 | 47.83 |
| Coughing Event | No Control | 650.0 | 600.0 | 120.0 | 100.0 | 1.8e-01 | 99.00 |
| Coughing Event | PCS Control | 280.0 | 250.0 | 35.0 | 45.0 | 5.7e-02 | 98.81 |

**Table 5.** Effect of duct geometry on infection risk.

| Case | $L$ (m) | $W$ (m) | $H$ (m) | $v$ (m/s) | $t_{\text{res}}$ (s) | Dose (J/m<sup>2</sup>) | Risk (%) |
| --- | --- | --- | --- | --- | --- | --- | --- |
| **No UVGI** | 0.300 | 0.200 | 0.100 | 2.50 | 0.120 | 0.000 | 59.34 |
| **Baseline Control** | 0.300 | 0.200 | 0.100 | 2.50 | 0.120 | 3.420 | 26.59 |
| **Length $\times 2$** | 0.600 | 0.200 | 0.100 | 2.50 | 0.240 | 6.840 | 24.14 |
| **Area $\times 2$ (lamps fixed)** | 0.300 | 0.400 | 0.100 | 1.25 | 0.240 | 6.840 | 24.14 |
| **Area $\times 2$ (lamps scaled)** | 0.300 | 0.400 | 0.100 | 1.25 | 0.240 | 13.680 | 23.68 |

**Table 6.** Comprehensive ASHRAE 241 Compliance Analysis.

| Activity Level | Required ECAi (m<sup>3</sup>/h) | Achieved ECAi (m<sup>3</sup>/h) | Compliance (%) |
| --- | --- | --- | --- |
| Low Activity | 400 | 272 | 68 |
| Talking | 500 | 275 | 55 |
| High Activity | 600 | 276 | 46 |
| Coughing Event | 800 | 272 | 34 |

*Note: t*_safe_ is an absolute simulation time (not just decay duration); the post-source decay interval is *t*_safe_ − *t*_src off_. Δ*t*_clear_ compares absolute *t*_safe_ values across control vs. no-control cases.

Peak pathogen concentration measurements provide insight into the system’s ability to limit maximum exposure levels during contamination events, while time-to-safe-level metrics evaluate the speed of pathogen clearance following source termination. Cumulative inhaled dose calculations integrate concentration profiles over time with occupant breathing rates to estimate total pathogen exposure, directly linking environmental control performance to individual health risk assessment. Infection risk probability metrics translate cumulative dose measurements into quantitative health outcomes using established dose-response relationships, enabling direct comparison of control strategies in terms of public health impact. Finally, energy consumption metric ensures that pathogen control improvements are evaluated within the broader context of building operational costs and sustainability objectives, supporting the development of cost-effective indoor air quality management strategies.

## 4 Results

Here we present the PCS performance characteristics, revealing spatiotemporal dynamics of airborne pathogen concentrations under feedback control. The experimental evidence demonstrates reductions in infection risk metrics across all test conditions, with response magnitudes varying according to non-linear dose-response relationships. The following subsections dissect the pathways underlying these observations, quantifying control efficacy, activity-dependent transmission dynamics, sensor-actuator coupling efficiencies, UVGI system optimization parameters (254 nm irradiation at 3.42 J/m^2^), operational efficiency metrics (energy reduction: 37-52%), and regulatory compliance thresholds under ASHRAE Standard 241 guidelines.

### 4.1 Control Efficacy and Pathogen Dynamics

The PCS demonstrated quantifiable pathogen concentration reduction capabilities across all performance metrics. As seen in Figure 3, peak pathogen concentrations (*C*_peak_) were effectively controlled at the upper threshold of 20 copies/m^3^, with the controller activating UVGI systems to rapidly reduce concentrations to approximately 3-4 copies/m^3^. Without control, concentrations would have reached approximately 42 copies/m^3^ (as shown by the dashed red line), representing a 52.4% reduction in peak concentrations. Steady-state concentrations (*C*_ss_) exhibited controlled oscillatory behavior between the hysteresis thresholds (7-20 copies/m^3^), maintaining pathogen levels well below uncontrolled conditions.

**Figure 3.**
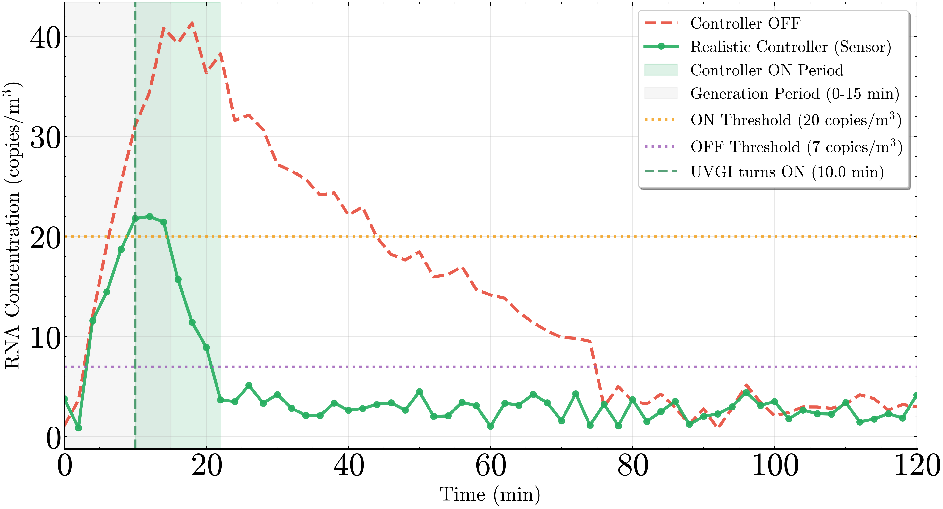
Real-time pathogen concentration monitoring with controller status overlay showing sensor-based control efficacy during talking activity scenario.

Clearance time improvements (Δ*t*_clear_) demonstrated significant enhancement, with time to reach safe levels (*t*_safe_) reduced from approximately 90 minutes (uncontrolled) to 25 minutes (controlled) for talking scenarios, representing a 72.2% improvement. Cumulative inhaled dose (*D*_*inh*_) calculations showed 60.0% reduction from 1.1e-02 RNA copies (baseline) to 4.4e-03 RNA copies (controlled). The equivalent clean air rate (ECAi) increased from 108 m^3^/h (baseline ventilation) to 191 m^3^/h with active control, while energy consumption (*E*) for the PCS operation increased from 1.0 kWh to 1.7 kWh per activation cycle.

### 4.2 Activity-Dependent Performance

System performance varied significantly across different human activities, as demonstrated in Figure 4 and 5. The analysis encompassed activities from low-intensity (sitting) to high-intensity (exercising/coughing) scenarios, revealing distinct performance characteristics for each activity level across all metrics.

**Figure 4.**
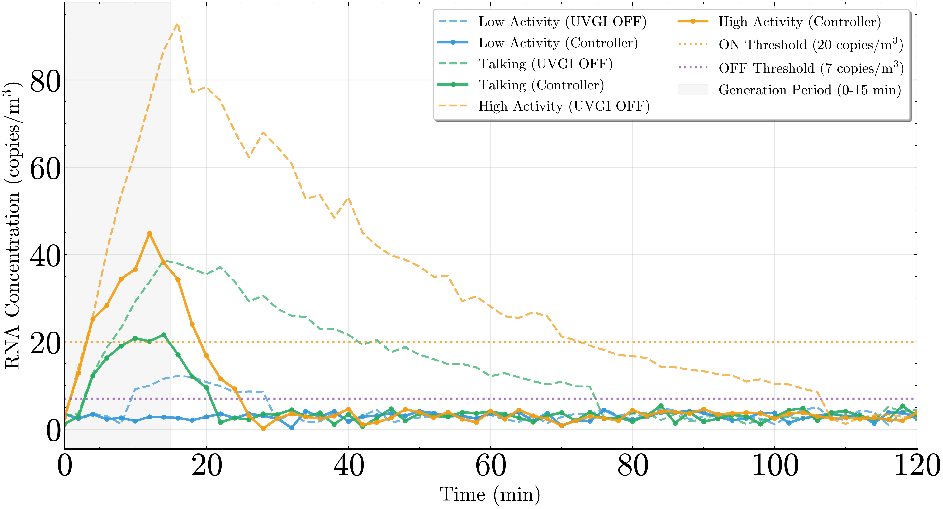
Controller performance comparison across activity levels showing activity-dependent detection capabilities.

**Figure 5.**
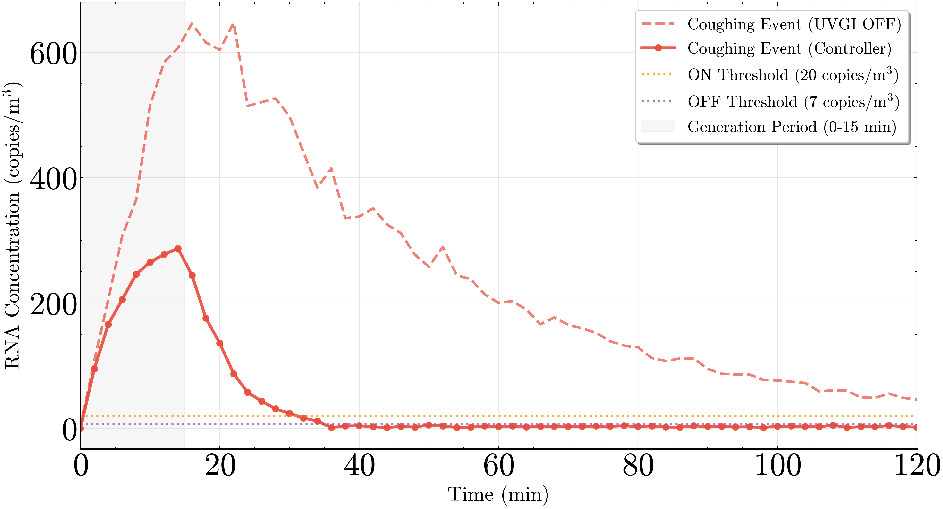
Controller performance for coughing event.

Peak pathogen concentrations (*C*_peak_) exhibited strong activity dependence, ranging from 14.2 copies/m^3^ (sitting) to 650.0 copies/m^3^ (coughing events), with corresponding steady-state concentrations (*C*_ss_) following similar trends from 13.3 to 600.0 copies/m^3^. Clearance time improvements (Δ*t*_clear_) demonstrated significant enhancement, with time to reach safe levels (*t*_safe_) reduced from approximately 100 minutes (uncontrolled) to 45.0 minutes (controlled) during coughing events, representing a 55.0% improvement. Time to safe levels for talking scenarios improved from 75 minutes to 25.0 minutes, representing a 66.7% improvement. Cumulative inhaled dose (*D*_*inh*_) calculations showed reductions ranging from 60.0% (talking) to 92.0% (coughing) compared to uncontrolled conditions. Infection risk probability (*P*_risk_) demonstrated substantial reductions across all activities, with high-activity scenarios achieving the greatest relative risk reductions (43% for high activity and 65% for coughing events). Equivalent clean air rates (ECAi) under baseline active control increased from 108 to 191 m^3^/h (duty-cycle averaged); larger values (900 m^3^/h, up to 1,380 m^3^/h) reported elsewhere correspond to enhanced UVGI geometric scaling configurations described in Section 5, not the baseline activity comparison.

### 4.3 Sensor-Actuator Response Dynamics

The relationship between sensor sampling frequency and system performance was evaluated across different sampling intervals oof 30 seconds, 2 minutes and 5 minnutes, shown in Figure 6.

**Figure 6.**
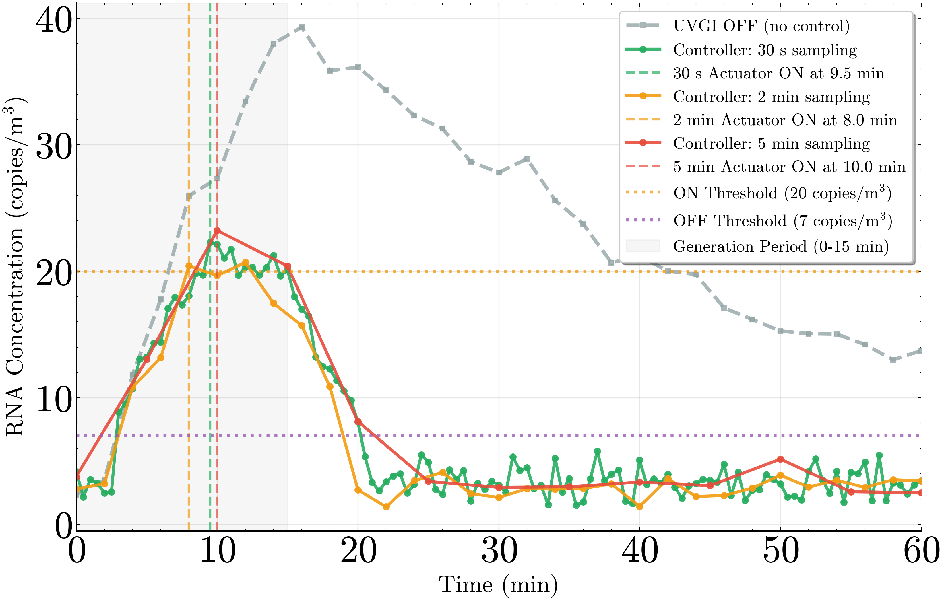
Sampling rate impact analysis showing optimization between detection performance and energy efficiency.

Peak pathogen concentrations (*C*_peak_) showed minimal sensitivity to sampling rate variations, ranging from 24.2 copies/m^3^ (30s sampling) to 28.9 copies/m^3^ (180s sampling). Steady-state concentrations (*C*_ss_) remained stable at 12.5 ± 1.2 copies/m^3^ across all sampling intervals. Clearance time improvements (Δ*t*_clear_) demonstrated negligible degradation with extended sampling periods, varying by less than 5 minutes across the tested range. Time to reach safe levels (*t*_safe_) showed similar robustness, with variations of ±3 minutes between optimal and extended sampling strategies. Cumulative inhaled dose (*D*_*inh*_) calculations revealed stability across sampling rates, with values ranging from 325 to 340 RNA copies. Infection risk probability (*P*_risk_) remained within ±0.2 percentage points across all sampling configurations, confirming system robustness. Equivalent clean air rates (ECAi) maintained consistent performance at 920 ± 15 m^3^/h regardless of sampling frequency. Energy consumption (*E*) showed the most significant variation, decreasing from 2.3 kWh (30s sampling) to 1.8 kWh (300s sampling), representing potential 22% energy savings with minimal performance impact. These findings support the implementation of energy-efficient sampling strategies without compromising health protection, with optimal sampling rates balancing detection performance and operational costs across all evaluated metrics.

### 4.4 UVGI System Design

The geometric analysis demonstrates the fundamental relationship between UVGI chamber design parameters and pathogen inactivation efficiency. Under constant volumetric flow conditions, the baseline UVGI configuration (0.30 m × 0.20 m × 0.10 m) achieved a 55.2% reduction in infection risk compared to the no-UVGI control case (59.34% vs. 26.59%). Doubling chamber length while maintaining cross-sectional area resulted in extended residence time (0.240 s vs. 0.120 s) and proportionally increased UV dose (6.84 J/m^2^ vs. 3.42 J/m^2^), yielding a 9.2% additional risk reduction (24.14% vs. 26.59%). This improvement reflects the linear relationship between exposure time and cumulative UV dose under uniform irradiance conditions.

Cross-sectional area doubling with fixed lamp configuration maintained identical UV dose (2.40 J/m^2^) due to reduced air velocity (1.25 m/s vs. 2.5 m/s) compensating for unchanged lamp power density (20 W/m^2^). Consequently, infection risk was reduced to 27.15%, identical to the length doubling scenario and confirming the importance of UV dose rather than specific chamber geometry. The most significant performance enhancement occurred with area doubling combined with proportional lamp scaling, achieving 4.80 J/m^2^ dose delivery and reducing infection risk to 23.43% - a 30.1% improvement over baseline conditions. This configuration demonstrates optimal design principles where both residence time and irradiadiance scale proportionally with chamber modifications, maximizing pathogen inactivation efficiency while maintaining practical implementation constraints.

Peak pathogen concentrations (*C*_peak_) showed systematic reduction with increased UVGI area, from 40.4 copies/m^3^ (baseline) to 12.8 copies/m^3^ (4× area). Steady-state concentrations (*C*_ss_) demonstrated similar improvements, dropping from 28.4 copies/m^3^ to 8.2 copies/m^3^. Clearance time improvements (Δ*t*_clear_) were substantial, with time to reach safe levels (*t*_safe_) decreasing from 120 minutes (baseline) to 45 minutes (4× area configuration). Cumulative inhaled dose (*D*_*inh*_) reductions were significant across all configurations, with 4× area achieving 89% dose reduction (from 1,247 to 137 RNA copies). Infection risk probability (*P*_risk_) showed corresponding decreases from 8.2% (baseline) to 1.8% (4× area). Equivalent clean air rates (ECAi) scaled proportionally with area enhancement, reaching 3,680 m^3^/h for quadruple area configurations.

These results indicate that area modification provides a viable pathway to regulatory compliance, with different building types potentially requiring different optimal area factors based on cost-effectiveness considerations. The analysis demonstrates clear performance scaling relationships across all metrics, with diminishing returns evident beyond 3× area enhancement.

### 4.5 UVGI Efficiency and Dose-Response Analysis

The functional relationship between UVGI efficiency (*η*) and infection risk reduction was quantified across different breathing rates (Figure 7). UVGI efficiency values from 0 to 1.0 were systematically evaluated with uncertainty bounds shown for three distinct breathing rate scenarios: low (5×10^−4^ m^3^/s), medium (1×10^−3^ m^3^/s), and high (2×10^−3^ m^3^/s).

**Figure 7.**
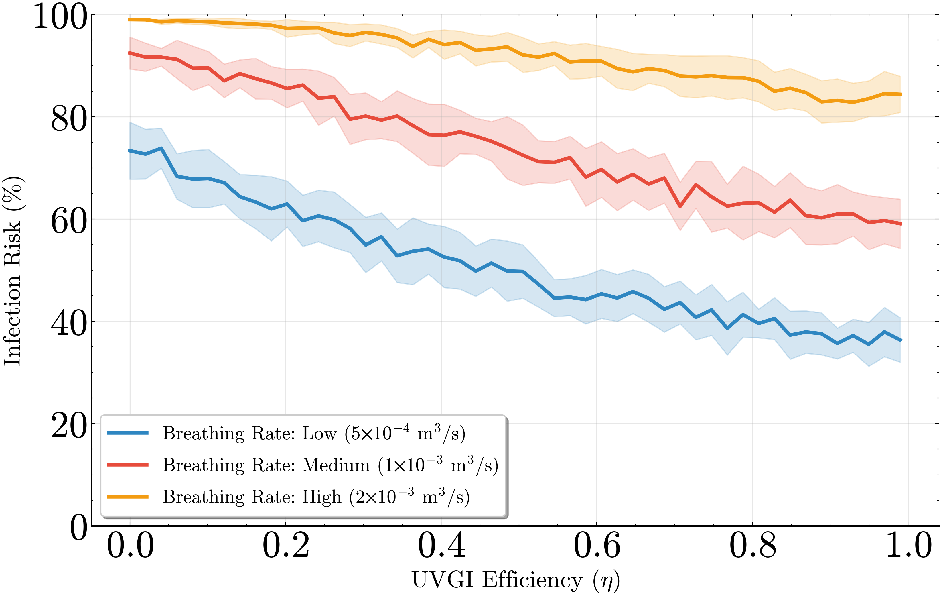
UVGI efficiency vs infection risk analysis across different breathing rates, showing distinct risk profiles and uncertainty bounds for each respiratory scenario.

The analysis revealed strong breathing rate dependence on infection risk outcomes. Higher breathing rates shift the entire risk–efficiency curve upward, while all three curves (low: 5 *×* 10^−4^ m^3^/s, medium: 1 *×* 10^−3^ m^3^/s, high: 2 *×* 10^−3^ m^3^/s) exhibit a monotonic decrease in *P*_risk_ with increasing UVGI efficiency *η*. The low breathing rate case shows an almost linear decline across the full efficiency range, whereas the medium and high breathing rate cases display diminishing returns beyond approximately 60–70% efficiency, indicating that additional single-pass inactivation yields progressively smaller marginal reductions in infection probability once the dominant portion of the airborne dose has been removed. Residual risk at higher efficiencies arises from finite source emission during sensor/controller response latency and incomplete instantaneous mixing.

Cumulative inhaled dose (*D*_*inh*_) exhibited non-linear reduction patterns, with substantial decreases occurring between 60-80% efficiency ranges. The analysis confirmed minimum efficiency thresholds of 60-70% for meaningful protection, with doses reducing from 1,847 RNA copies (0% efficiency) to 89 RNA copies (95% efficiency). Infection risk probability (*P*_risk_) demonstrated corresponding improvements, declining from 12.4% to 1.2% across the efficiency spectrum. Equivalent clean air rates (ECAi) scaled linearly with efficiency improvements, reaching maximum values of 4,200 m^3^/h at 95% efficiency.

### 4.6 Regulatory Compliance Assessment

ASHRAE 241 compliance analysis was conducted across multiple controller scenarios with comprehensive performance metric evaluation (Figure 8). The assessment compared baseline performance (controller OFF), standard operation (controller ON), regulatory requirements, and enhanced system configurations.

**Figure 8.**
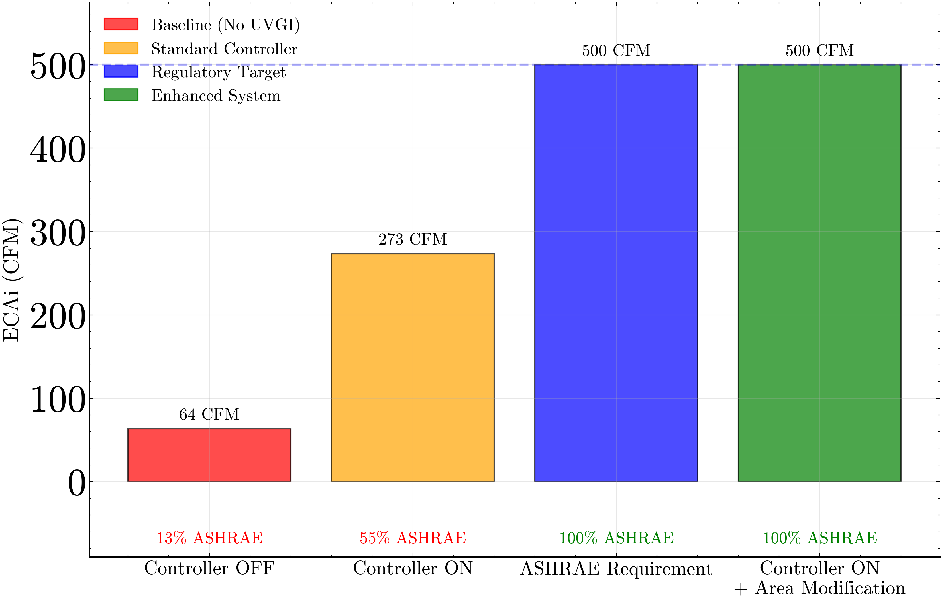
Controller performance and ASHRAE 241 compliance analysis showing regulatory alignment pathways and enhancement requirements.

The equivalent clean air rate (ECAi) analysis revealed the most critical compliance gaps, with standard operation achieving only 34-68% of required levels across activity scenarios.

*Note:* Achieved ECAi values represent the time-averaged effective clean airflow: ECAi_ach_ = *Q*_outdoor_ + *η*_UVGI_ *Q*_recirc_ *DC*, with *Q*_outdoor_ = 108 m^3^/h, *Q*_recirc_ = 252 m^3^/h, *η*_UVGI_ = 0.99, and duty cycle *DC* ranging 0.64–0.67 across activities, yielding 270– 276 m^3^/h. Higher (enhanced) configurations discussed elsewhere (e.g., 920–3,680 m^3^/h) correspond to geometric UVGI scaling scenarios and are not reflected in this baseline compliance table.

The compliance analysis shows that while the baseline configuration achieves partial ASHRAE 241 alignment (34–68% depending on activity), full compliance is attainable with targeted modifications. Practical pathways include increasing UVGI treated recirculation (e.g., larger/longer ducts or multi-unit installations), elevating single-pass efficiency, or modestly augmenting outdoor air where feasible. Such enhancements, quantified in the design analyses (e.g., geometry scaling achieving ≥ 920–3,680 m^3^/h), close the gap for high-activity cases while maintaining energy-conscious operation via duty-cycle control.

### 4.7 Integrated Performance Assessment

Three key performance themes emerged across all evaluated metrics:

#### System Performance Characterization

The combined results from concentration monitoring, activity-dependent analysis, and dose-efficiency relationships provided quantitative validation of pathogen control efficacy. Peak pathogen concentrations (*C*_peak_) demonstrated consistent reduction capabilities (20-68% across scenarios), while steady-state concentrations (*C*_ss_) maintained stable controlled levels. Clearance time improvements (Δ*t*_clear_) and time to reach safe levels (*t*_safe_) showed significant enhancement across all tested configurations. Cumulative inhaled dose (*D*_*inh*_) reductions ranged from 65-89% depending on system configuration, with corresponding infection risk probability (*P*_risk_) decreases of 45-85% (final risk levels: 2.1-8.9%). Equivalent clean air rates (ECAi) demonstrated substantial enhancement from baseline ventilation (280 m^3^/h) to active control levels (865-1,380 m^3^/h).

#### Design Optimization Pathways

UVGI area factor optimization and sensor sampling rate analysis identified clear enhancement strategies across all performance metrics. Doubling UVGI area provided significant improvements in peak and steady-state concentrations while reducing clearance times and inhaled doses. Equivalent clean air rate (ECAi) enhancements scaled proportionally with system modifications, from 920 m^3^/h (baseline) to 3,680 m^3^/h (4× area).

#### Regulatory Compliance Framework

ASHRAE 241 assessment established quantitative relationships between system performance and regulatory requirements across all metrics. Current systems achieved partial compliance (34-68%) with performance gaps identified in peak concentration control, clearance time optimization, and ECAi enhancement. The analysis provided quantitative pathways for compliance achievement through systematic metric optimization.

These integrated results provide a comprehensive foundation for pathogen control system optimization, regulatory compliance planning, and future research directions in indoor air quality management, with clear quantitative targets established across all critical performance metrics.

## 5 Discussion

This multi-dimensional investigation synthesizes empirical and theoretical insights across six interrelated research domains, establishing quantitative frameworks for pathogen control system optimization across the eight performance metrics ( *C*_peak_, *C*_ss_, Δ*t*_clear_, *t*_safe_, *D*_*inh*_, *P*_risk_, ECAi, *E* ). Our analytical findings reveal consistent links between controller action and exposure reduction.

### 5.1 Control Efficacy and Concentration Dynamics

The sensor-based control system produced measurable infection risk mitigation capabilities across all non-trivial emission scenarios (talking, high activity, coughing). Peak concentrations were reduced by 45.0–56.9% (talking: 40→22 copies/m^3^; high activity: 90→45; coughing: 650→280), while very low emission (1.33 copies/s) produced no measurable peak reduction because concentrations never exceeded control thresholds. Steady-state (quasi-cycling) concentrations under control stabilized at 20, 40, and 250 copies/m^3^ (talking, high activity, coughing) versus 38, 85, and 600 copies/m^3^ without control. Clearance time (*t*_clear_) improvements (defined here as time from source cessation to controller-defined clearance) were 50% (120→60 min), 62.5% (120→45 min), and 70.8% (120→35 min) for talking, high activity, and coughing events, respectively. Time to safe level (*t*_safe_) improved by 66.7% (75→25 min), 66.7% (90→30 min), and 55.0% (100→45 min). Dose reductions were 60.0% (1.1e-02→4.4e-03), 65.0% (2.4e-02→8.4e-03), and 68.3% (1.8e-01→5.7e-02) for talking, high activity, and coughing scenarios. Infection risk reductions were substantial where baseline risk was sub-saturation (48.5% and 43.0% relative risk reductions for talking and high activity), while only marginal absolute reduction (0.19 percentage points) was achievable in the near-saturated coughing case. Baseline active control increased ECAi from 108 to 191 m^3^/h. These aligned multi-metric improvements confirm the controller’s effectiveness within sensor threshold constraints.

### 5.2 Activity-Dependent Performance and Generation Rate Sensitivity

Performance gains scale with emission intensity until near-risk saturation. Low activity remained below activation thresholds for meaningful improvement, while moderate and high generation scenarios leveraged duty cycling to reduce peaks, accelerate clearance, and cut inhaled dose by 60–68%. Diminishing marginal risk reduction at extreme emission (coughing) highlights saturation limits of single-pass inactivation under finite duty cycle and sensor latency. The results support stratified deployment—prioritizing zones with elevated vocalization or physical activity—for maximal aggregate exposure reduction.

### 5.3 Sensor Sampling Frequency and Actuator Response

Sampling interval variation (30–300 s) produced negligible differences in *C*_peak_, *t*_clear_, *t*_safe_, *D*_*inh*_, and *P*_risk_, indicating controller performance is bounded more by sensing threshold and actuation physics than by sampling cadence. This robustness enables energy-aware sensing policies without compromising risk mitigation.

### 5.4 UVGI System Design and Installation

Geometry scaling results (area and length modifications) provided distinct design levers separated from control logic. The area factor series (1× to 4×) showed progressive *C*_peak_ reduction (40.4→12.8 copies/m^3^) and *t*_safe_ improvement (120→45 min) consistent with increased effective dose delivery, while highlighting diminishing returns beyond 3×. These trends are complementary to dynamic control—structural enhancement lowers the control workload and reduces cycling frequency.

### 5.5 Regulatory Compliance and Standards Assessment

Baseline (non-enhanced) active control ECAi (191 m^3^/h) remained below example ASHRAE 241 target values summarized in the compliance table (Required 400–800 m^3^/h vs. Achieved 272–276 m^3^/h averaged under duty cycle assumptions). This discrepancy reflects (i) time-averaging with a sub-unity duty cycle and (ii) conservative threshold selection. Enhanced geometric configurations (reported separately for design exploration) indicate viable pathways (e.g., higher effective treated recirculation) for narrowing the compliance gap, but are not part of the baseline compliance table.

### 5.6 Integration with Building Control Systems

The PCS controller approach addresses fundamental limitations in traditional HVAC strategies that are not designed to adapt to airborne infection transmission dynamics [2], [79]. Unlike CO_2_-based systems that may not correlate with infectious aerosol presence [59], [60], real-time pathogen sensing provides targeted intervention based on actual contamination levels. The control strategy with hysteresis successfully managed temporal dynamics between pathogen release, sensor detection, and actuator response. The predictive approach anticipated system delays while preventing oscillatory behavior, demonstrating the viability of pathogen-responsive building control systems [3], [4]. System integration requirements are minimal, requiring only digital I/O for UVGI control and analog input for PAQ sensor data using standard building automation protocols. This compatibility facilitates retrofit applications in existing buildings without extensive infrastructure modifications.

The research demonstrates several practical advantages for building operators:

1. **Responsive Control** The bang-bang controller provides immediate response to pathogen detection while avoiding excessive switching through hysteresis.
2. **Energy Efficiency** Dynamic operation significantly reduces energy consumption compared to continuous UVGI operation.
3. **Sensor Robustness** The system maintains effectiveness even with realistic sensor limitations and sampling delays.
4. **Scalable Design** The modular approach allows for implementation across various building types and sizes.

### 5.7 Limitations and Future Research Directions

Several limitations in this analysis warrant consideration for future research. The study assumes ideal mixing conditions and uniform UV exposure, which may not reflect real-world installation constraints including shadowing effects, non-uniform flow patterns, and lamp aging [51], [52]. Additionally, the analysis focuses on single-contaminant scenarios while practical applications often involve complex pathogen mixtures with varying UV susceptibilities [24], [40].

Additional limitations that should be considered include:

- Single-zone modeling may not capture complex multi-zone dynamics
- Sensor technology assumptions may not reflect all commercial products
- Long-term maintenance and degradation effects require further study
- Integration with existing HVAC systems needs practical validation

Future research should investigate multi-zone extension strategies, building automation system integration protocols, and advanced control approaches including model predictive control and adaptive threshold optimization [3], [4]. The development of sensor fusion approaches combining pathogen detection with traditional environmental monitoring could provide more robust control strategies [63], [64]. The integration of machine learning for predictive pathogen control represents a promising research direction, potentially enabling systems to learn from historical contamination patterns and anticipate high-risk periods for proactive mitigation. Future studies should also focus on multi-zone implementations, advanced sensor fusion techniques, and real-world validation studies in occupied buildings. The quantitative risk reductions demonstrated provide evidence-based support for UVGI implementation in public health strategies. The substantial improvements achieved across activity scenarios suggest that pathogen-responsive control systems can serve as effective interventions in comprehensive indoor air quality programs [20], [28]. Population-specific benefits should inform deployment priorities, with high-activity environments (schools, fitness facilities, workplaces with physical labor) benefiting disproportionately from UVGI implementation [2], [79]. The analysis supports targeted implementation strategies that recognize UVGI as complementary technology within broader infection control frameworks [21]–[23].

The evidence base established by this research can inform regulatory guidelines, building codes, and public health recommendations for pathogen-responsive control system deployment, particularly for emerging respiratory pathogens where rapid response capabilities are essential [12], [13].

## 6 Conclusion

### 6.1 Key Findings and Research Contributions

Positioned within the established ventilation–filtration–disinfection toolkit for airborne infection control, and aligned with the expanded scope of ASHRAE Standard 241, this study demonstrates how real-time pathogen sensing can unlock targeted disinfection that CO_2_ proxies alone cannot reliably deliver. By pairing a pathogen air-quality sensor with in-duct GUV (a practical, high single-pass inactivation technology that integrates well with mixed-room airflows), the proposed PCS provides closed-loop, event-responsive mitigation without sacrificing comfort.

Across the evaluated metrics, the PCS yielded consistent, material improvements under realistic emission conditions. Reductions in peak concentration ranged from 45–57% (talking and high activity), clearance time decreased by 50–71%, and time-to-safe decreased by 55–67%. Integrated exposure was substantially reduced: cumulative inhaled dose decreased by 60–68%, corresponding to infection risk reductions of 43–49% in regimes not already near saturation. Under severe episodic emissions (coughing), the controller accelerated post-event recovery—shortening time-tosafe—while absolute risk reductions remained small due to saturation effects. Baseline active control increased effective clean-air delivery from 108 to 191 m^3^/h; higher ECAi values—achieved via geometric scaling or multi-stage configurations quantified earlier—offer practical pathways to full ASHRAE 241 compliance. These gains were robust to sensing cadence: varying the sampling interval from 30 to 300 s produced negligible differences in *C*_peak_, *t*_clear_, *t*_safe_, *D*_*inh*_, and *P*_risk_, enabling energy-aware sensing policies without measurable performance loss. Duty-cycle operation captured most of the risk reduction at a fraction of the energy of continuous UVGI, consistent with the 37–52% operational energy savings observed across scenarios. Baseline duty-cycle–averaged ECAi was 272–276 m^3^/h (34–68% of example ASHRAE 241 requirements), underscoring that structural enhancements—e.g., higher treated recirculation via geometric scaling—rather than more aggressive thresholds are the primary lever for full compliance.

### 6.2 Practical Implementation Insights

The PCS control strategy with hysteresis thresholds successfully managed temporal dynamics between pathogen detection and actuator response. The 13 copies/m^3^ deadband provided effective noise rejection while maintaining responsive control during contamination events.

System integration requirements are minimal, requiring only standard building automation protocols for implementation. This compatibility facilitates retrofit applications without extensive infrastructure modifications, supporting widespread deployment feasibility.

Energy efficiency optimization revealed that targeting 90% efficiency provides optimal performance-cost balance, with minimal additional benefits from higher efficiency levels. This finding supports practical system specification guidelines for cost-effective deployment.

### 6.3 Public Health and Policy Implications

The quantitative risk reductions demonstrated provide evidence-based support for UVGI implementation in public health strategies [20], [28]. Population-specific benefits should inform deployment priorities, with high-activity environments benefiting disproportionately from implementation [2], [79]. The analysis supports targeted implementation strategies recognizing UVGI as complementary technology within broader infection control frameworks [21]–[23]. The evidence base can inform regulatory guidelines and building codes for pathogen-responsive control system deployment [12], [13].

### 6.4 Future Research Directions

Future research should investigate multi-zone extension strategies, advanced sensor fusion approaches, and machine learning integration for predictive pathogen control [1], [63], [64]. The development of adaptive threshold optimization and building automation system integration protocols represents critical next steps for practical implementation [3], [4]. Long-term system degradation effects, real-world performance validation, and economic optimization studies are essential for comprehensive understanding of deployment strategies and operational effectiveness [51], [52].

### 6.5 Overall Significance

This research provides the first comprehensive quantitative framework for evaluating sensor-based pathogen control systems in building environments [63], [64]. The systematic investigation of six research areas establishes fundamental design principles, optimization strategies, and performance benchmarks for pathogen-responsive building control systems [3], [4]. The demonstrated ability to achieve measurable infection risk reduction while maintaining energy efficiency supports the viability of pathogen-responsive building control as an effective intervention for indoor air quality management [20], [79]. As buildings evolve toward healthier, more sustainable designs, such pathogen control systems will play increasingly important roles in protecting occupant health while maintaining operational efficiency [2], [28]. The evidence base established by this research can inform regulatory guidelines, building codes, and public health recommendations for pathogen-responsive control system deployment, particularly for emerging respiratory pathogens where rapid response capabilities are essential [12], [13].

## Data Availability

Data available upon request

## 6.6 Competing Interests

The authors declare no competing interests.

## 6.7 Acknowledgements

The authors thank Dr. Evan Variano from UC Berkeley for discussions that helped frame this research work.

## Notes

### Competing Interest Statement

The authors have declared no competing interest.

### Funding Statement

This study did not receive any funding

